# Combined administration of inhaled DNase, baricitinib and tocilizumab as rescue treatment in COVID-19 patients with severe respiratory failure

**DOI:** 10.1101/2022.03.14.22270915

**Authors:** Efstratios Gavriilidis, Christina Antoniadou, Akrivi Chrysanthopoulou, Maria Ntinopoulou, Andreas Smyrlis, Iliana Fotiadou, Nikoleta Zioga, Dionysios Kogias, Anastasia-Maria Natsi, Christos Pelekoudas, Evangelia Satiridou, Stefania-Aspasia Bakola, Charalampos Papagoras, Ioannis Mitroulis, Paschalis Peichamperis, Dimitrios Mikroulis, Vasileios Papadopoulos, Panagiotis Skendros, Konstantinos Ritis

## Abstract

COVID-19-related severe respiratory failure (SRF) leads to mechanical ventilation increasing the in-hospital mortality substantially. Abundancy of lung fibroblasts (LFs) in injured lung tissue has been associated with the progression of respiratory failure in COVID-19. Aiming to reduce mortality in patients with SRF (PaO2/FiO2<100 mmHg) and considering the multi-mechanistic nature of severe COVID-19 pathogenesis, we applied a combined rescue treatment (COMBI) on top of standard-of-care (SOC: dexamethasone and heparin) comprised inhaled DNase to dissolve thrombogenic neutrophil extracellular traps, plus agents against cytokine-mediated hyperinflammation, such as anti-IL-6 receptor tocilizumab and selective JAK1/2 inhibitor baricitinib. COMBI (n=22) was compared with SOC (n= 26), and with two previously and consecutively used therapeutic approaches, including either IL-1 receptor antagonist anakinra (ANA, n=19), or tocilizumab (TOCI, n=11), on top of SOC. In parallel, evaluation of immunothrombosis was assessed in vitro in human LFs, treated with the applied therapeutic agents upon stimulation with COVID-19 plasma. COMBI was associated with lower in-hospital mortality (p=0.014) and intubation rate (p=0.013), shorter duration of hospitalization (p=0.019), and prolonged overall survival after a median follow-up of 110±4 days (p=0.003). In vitro, COVID-19 plasma markedly induced tissue factor/thrombin pathway in LFs, while this effect was inhibited by the immunomodulatory agents of COMBI providing a mechanistic explanation for the clinical observations. These results suggest the design of randomized trials using combined immunomodulatory therapies in COVID-19-associated SRF targeting multiple interconnected pathways of immunothrombosis.

## 1. Introduction

In patients with COVID-19, severe hypoxemic respiratory failure (SRF) leading to invasive mechanical ventilation (IMV), raises the mortality rate substantially [1–3]. Thus, the management of patients with SRF to avoid intubation and intensive care admission is a challenging and crucial issue [2–4]. Numerous clinical trials were conducted or are underway in the efforts to improve the current mainstay of treatment for patients with severe COVID-19, which consists of dexamethasone, heparin, and supportive care [5,6].

Deterioration of COVID-19 respiratory function seems to be associated with progressed dysregulation of immune responses [7]. Severe acute respiratory syndrome coronavirus 2 (SARS-CoV-2) infects predominantly alveolar epithelial cells, triggering aberrant inflammatory responses leading to acute respiratory distress syndrome (ARDS) [8,9]. Neutrophil extracellular traps (NETs) are related with the activity of various immunological mediators and the subsequent inflammatory milieu [10]. Moreover, advanced respiratory disease in COVID-19 is characterized by macrophage extravasation, while an abundance of mesenchymal cells and fibroblasts is observed and linked with the progression of respiratory failure [8,9].

Mechanistically, NET-mediated immunothrombosis, leads to activation of tissue factor (TF) axis and thrombin generation. Proteases of this axis fuel a vicious cycle, in which a harmful crosstalk between thrombosis, inflammation and fibrosis is developed [8,11]. Moreover, complement activation, triggering of the NF-κB and JAK/STAT pathways, have been described as partners in COVID-19 hyperinflammation [11–13]. Based on these mechanisms and the current ongoing randomized control trials (RCTs), immunomodulatory treatments including anakinra, a recombinant IL-1 receptor antagonist [14], tocilizumab, an anti-IL-6 receptor antagonist [15], baricitinib, a selective JAK-1/JAK-2 inhibitor [16] and nebulized recombinant human DNase [17,18] as an agent which dismantles the generated NETs, have been administered separately in patients with COVID-19-related SRF.

This study describes, as rescue treatment, a therapeutic protocol based on the multi-mechanistic nature of severe COVID-19. Standard-of-care (SOC) in conjunction with a combined therapeutic strategy, comprising inhaled DNase plus baricitinib and tocilizumab, was found to reduce the mortality in a group of patients with severe respiratory failure. Translating the clinical findings, we observed diminished expression of TF/thrombin axis in primary lung fibroblasts treated with the applied therapeutic agents upon stimulation with COVID-19 environment.

## 2. Methods

### 2.1 Study design and patients

#### 2.1.1 Criteria of the study

This is a non-randomized open-label study, conducted in the First Department of Internal Medicine, University Hospital of Alexandroupolis. In total, 368 patients with COVID-19 were consecutively admitted from October 25, 2020 to August 18, 2021. Among them, we studied 78 patients who progressed to severe respiratory failure (SRF) defined as a ratio of arterial partial pressure of oxygen to fraction of inspired oxygen (PaO2/FiO2) <100 mmHg [2]. They received SOC treatment at the time of admission or combination of SOC with known immunomodulating factors, in a consecutive chronologically pattern, as compassionate/rescue treatment (**Figure 1**).

**Figure 1.**
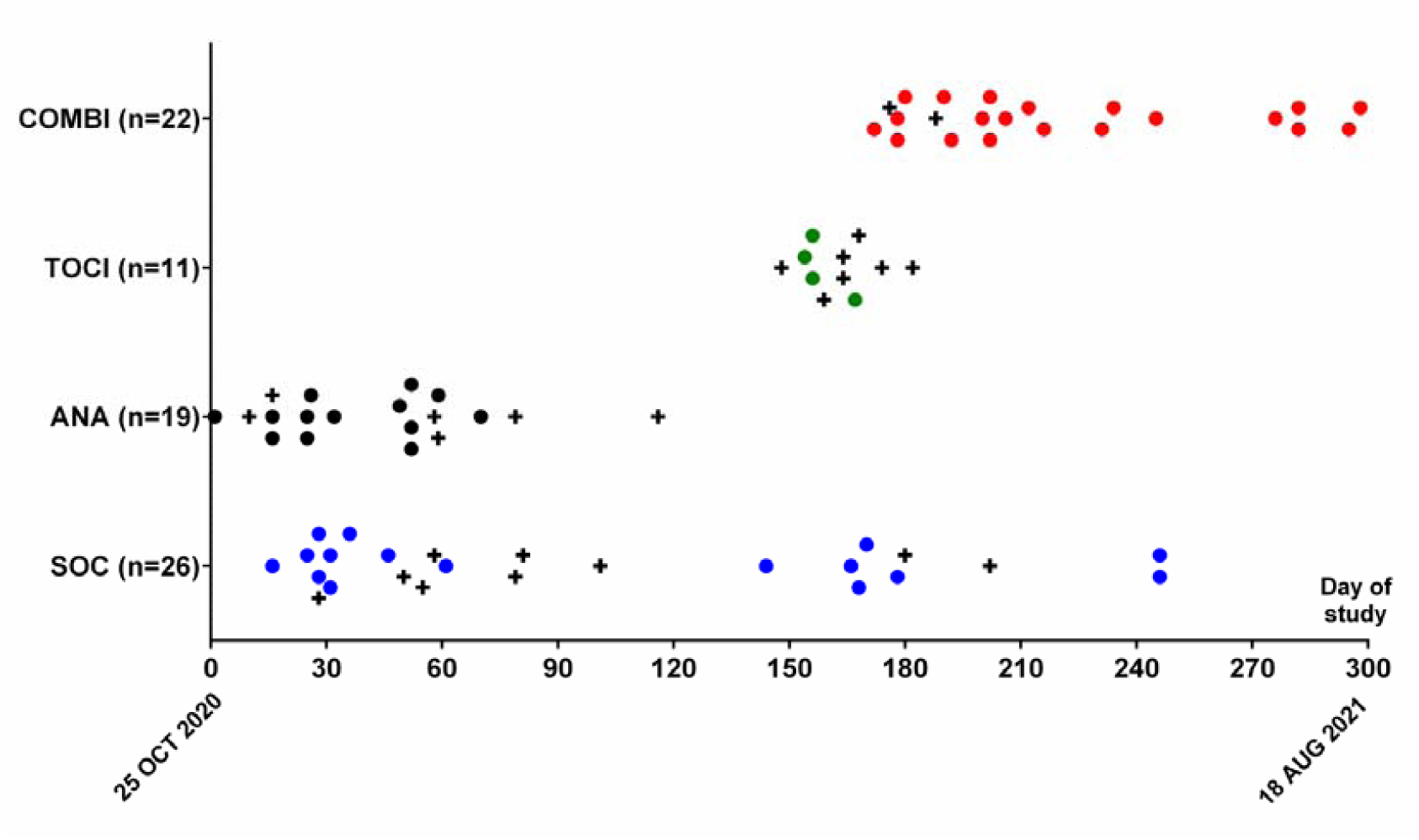
Consecutive groups of therapies and number (n) of patients enrolled during the study period; the symbol + denotes in-hospital deaths.

Inclusion criteria were as follows: 1) adult patients ≥18 years old, of any gender, 2) positive polymerase-chain-reaction (PCR) test for SARS-CoV-2 RNA in nasopharyngeal swab, 3) pulmonary infiltrates suggestive of COVID-19, 4) SRF as defined by PaO2/FiO2<100 mmHg, 4) written informed consent from the patients or legal representatives for the current compassionate therapeutic protocol.

Patients were excluded from the analysis according to the following criteria: 1) need for intubation/IMV during the first 24 hours after the initiation of treatment, 2) multi-organ failure, 3) systemic co-infection, 4) SRF due to cardiac failure or fluid overload, 5) glomerular filtration rate (GFR) <30 ml/min/1.73 m^2^), 6) any stage IV solid tumor or immunosuppression due to hematological disorders, 7) any immunosuppressive therapy and/or chemotherapy during the last 30 days, 8) low patient’s functional performance as defined by a Palliative Performance Scale (PPS) score ≤30% [19], 9) pregnancy.

#### 2.1.2 Study groups and endpoints

The selection of the different rescue treatments that have been used throughout the study period, was initially decided, and periodically revised, by considering any new published or ongoing RCT, as well as emerging research findings of our group and others on the role of multiple pathways in the inflammatory milieu of severe COVID-19, including the critical role of NETs, the triggering of NFκB pathway and the induction of IL-6 and Janus kinases (JAKs). In total, 4 SRF groups were compared and analyzed, including the group of patients treated with SOC only and 3 consecutive chronologically additional groups in which apart from SOC treatment, different rescue therapeutic immunomodulatory regimens have been added (**Figure 1**).

SOC group comprised patients who received only standard of care in accordance to our department protocol including dexamethasone 6-8 mg once daily, low molecular weight heparin at therapeutic doses, antibiotic prophylaxis and supportive care.

TOCI group included patients who received SOC plus IL-6 receptor inhibitor tocilizumab. Tocilizumab was administered as a single intravenous (iv) dose of 8 mg/kg actual body weight up to 800 mg.

ANA group included patients who received SOC plus IL-1 receptor antagonist anakinra. Anakinra was administered iv 200 mg/twice daily for 3-6 days, then 100 mg/twice daily, for up to 10 days in total.

COMBI group included patients who were treated with SOC and the combination following regimens: a) tocilizumab (as described above), b) selective JAK-1/JAK-2 inhibitor baricitinib, 4 mg per os once daily, for up to 14 days (2 mg if GFR was 30 to <60 ml/min/1.73 m^2^) and c) tο dismantle the accumulated NETs in lung, nebulized dornase alfa, a recombinant human DNase I (inhaled DNase), 2,500 U/twice daily, for up to 14 days. Inhaled DNase was administered simultaneously with inhaled budesonide (800 µg/twice daily) and bronchodilators (salbutamol or/and ipratropium at conventional doses).

During the course of the study, the same general care protocols were used by the nursing personnel. Baseline demographical and clinical characteristics of the patients are demonstrated in **Table 1**.

**Table 1.** Baseline demographical and clinical characteristics of the patients studied (n=78).

| Parameter | SOC<br>n=26 | ANA<br>n=19 | TOCI<br>n=11 | COMBI<br>n=22 | *p* |
| --- | --- | --- | --- | --- | --- |
| **Age (years)** |  |  |  |  |  |
| Mean±SD | 62.8±10.6 | 62.9±10.9 | 63.6±8.7 | 56.9±11.1 | 0.154 |
| **Sex (%)** |  |  |  |  |  |
| Male (%) | 18 (69.2) | 15 (78.9) | 8 (72.7) | 16 (72.7) | 0.912 |
| Female (%) | 8 (30.8) | 4 (21.1) | 3 (27.3) | 6 (27.3) |  |
| **Body Mass Index (BMI)** |  |  |  |  |  |
| Mean±SD | 30.3±4.0 | 29.7±5.3 | 29.8±4.7 | 30.9±5.1 | 0.853 |
| **Number of comorbidities** |  |  |  |  |  |
| Mean±SD | 2.2±1.4 | 2.0±1.4 | 2.0±1.1 | 1.8±1.1 | 0.722 |
| 0 (%) | 3 (11.5) | 4 (21.1) | 1 (9.1) | 2 (9.1) |  |
| 1 (%) | 6 (23.1) | 3 (15.8) | 3 (27.3) | 8 (36.4) |  |
| 2 (%) | 6 (23.1) | 5 (26.3) | 2 (18.2) | 6 (27.3) |  |
| 3 (%) | 6 (23.1) | 4 (21.1) | 5 (45.4) | 5 (22.7) |  |
| 4 (%) | 4 (15.4) | 3 (15.8) | 0 (0) | 1 (4.5) |  |
| 5 (%) | 1 (3.8) | 0 (0) | 0 (0) | 0 (0) |  |
| **Disease day at admission\*** |  |  |  |  |  |
| Mean±SD | 9.5±2.9 | 9.3±3.9 | 7.7±3.1 | 9.5±1.8 | 0.361 |
| **Disease day at enrollment** |  |  |  |  |  |
| Mean±SD | 11.3±2.6 | 11.6±3.1 | 10.0±2.4 | 10.6±2.2 | 0.358 |
| **PaO<sub>2</sub>/FiO<sub>2</sub> at enrollment (mmHg)** |  |  |  |  |  |
| Mean±SD | 80.9±12.5 | 89.8±29.2 | 87.9±24.9 | 96.0±21.6 | 0.129 |
| **SOC treatment** |  |  |  |  |  |
| Dexamethasone | 26 | 19 | 11 | 22 | 1.000 |
| Low molecular weight heparin | 26 | 19 | 11 | 22 |  |
| Antibiotics | 26 | 19 | 11 | 22 |  |

\* The onset of the disease was defined as the date of the first symptoms consistent with COVID-19 or, whenever this was not feasible, the date of the first positive PCR.

Primary endpoint was the reduction of the in-hospital mortality rate, whereas secondary endpoints concerned intubation/IMV rate, days of hospitalization and overall survival as derived from the last follow-up visit, either in-office or remote.

The study was aligned with the Helsinki Declaration. Subjects provided written informed consent before study participation. Patients’ records were anonymized and deidentified prior to analysis and so confidentiality and anonymity were assured. The study protocol design was approved by the local scientific and ethics committees and institutional review board of the University Hospital of Alexandroupolis (Ref. No. 87/08-04-2020).

### 2.2 Statistical Analysis

#### 2.2.1 Analysis of clinical data

Chi square test along with adjusted residuals was used to compare outcomes among the four treatment groups. Similarly, chi square and ANOVA were used to compare binary and discrete/continuous variables, respectively, that are considered to be potential confounders. To consolidate the independent correlation of each treatment group with outcome, a Generalized Linear Model using outcome as dependent variable, treatment groups as independent variables and potential confounders as factors was further utilized. For that purpose, all scale variables were turned into binary ones with the use of Optimal Scaling along with ridge regression, random initial configuration and bootstrapping. Secondary outcomes were evaluated with ANOVA; ad-hoc analysis was performed using Tuckey’s HSD test. The repeated measures General Linear Model was used for analysis of within-subjects and between-subjects variance of the same variable measured several times on each patient. Kaplan-Meier curves were used to depict survival data; comparisons were performed by the Logrank test. A Cox proportional-hazards regression model was introduced to examine simultaneously the effects of multiple covariates on overall survival. Median follow-up was approached by the reverse Kaplan-Meier estimator.

#### 2.2.2 Analysis of experimental data

Comparisons between two groups were performed using Student’s t test (2-tailed). For comparisons among more than two groups, Kruskal-Wallis test, followed by Dunn’s test for multiple comparisons, was performed.

For both clinical and experimental results, the level of statistical significance was set at 0.05. Bonferroni correction was applied in case of multiple testing. Data are presented as mean ± standard deviation (SD). Statistical analysis was performed using SPSS 26.0 software.

Detailed information for all methods can be found in the in the Supplementary Materials section.

## 3. Results

### 3.1 Clinical outcomes

Analysis of baseline characteristics revealed that all groups of SRF patients were comparable at enrollment (**Table 1**).

In the SOC group, 9 deaths were recorded among 26 patients (34.6%). 10 out of 26 patients were intubated (38.5%); 8 of them did not survive. The mean duration of hospitalization was 19.4 days (SD: ±7.2). In the ANA group, 6 deaths and 6 intubations were recorded among 19 hospitalized patients (31.6%). Five intubated patients did not survive. The mean duration of hospitalization was 23.9 days (SD: ±10.8). In the TOCI group, 7 deaths were recorded out of 11 patients, all who had been intubated (63.6%). The mean duration of hospitalization was 19.4 (SD: ±7.8). In the COMBI group, there were 2 deaths among 22 patients (9.1%), both in intubated patients. There was a reduction in the mean duration of hospitalization, which was 15.6 days (SD: ±5.3).

Taken together, COMBI treatment was associated with significant lower in-hospital mortality (p=0.014), as well as intubation rate (p=0.013) and shorter duration of hospitalization (p=0.019) compared to other therapeutic groups of patients with COVID-related SRF (**Figure 2A**,**B**). Moreover, COMBI treatment was correlated with prolonged survival (p=0.003) after a median follow-up of 110±4 days; loss-of-follow-up was 1.3% (1 patient at SOC group) (**Figure 2C**).

**Figure 2.**
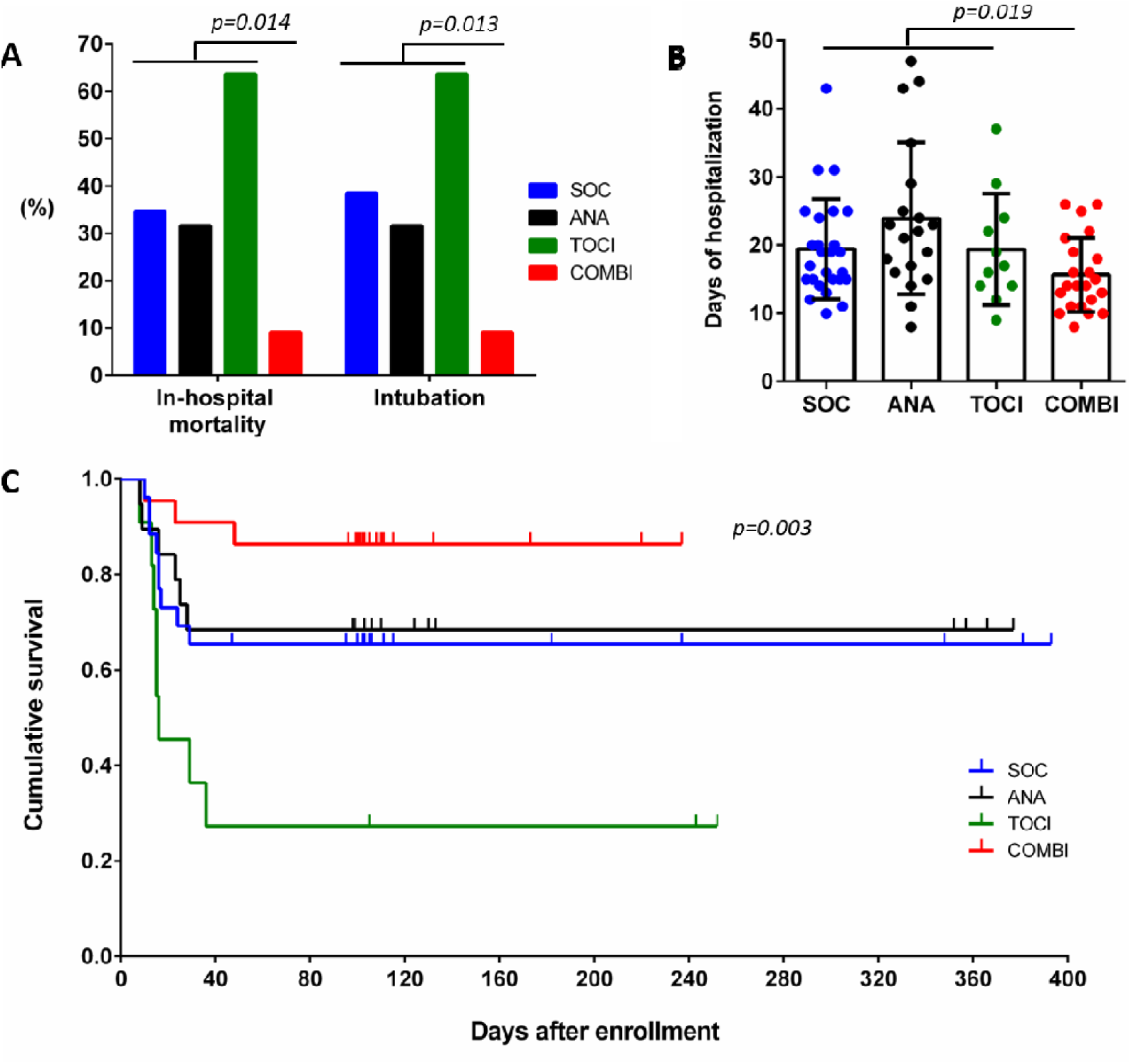
COMBI treatment reduces **(A)** mortality and intubation rate, and **(B)** days of hospitalization, whereas **(C)** prolongs overall survival when compared with other treatments.

A Generalized Linear Model was used to assess the effect of potential confounders in mortality rate; similarly, a Cox regression proportional hazards model, incorporating the same potential confounders as covariates, was introduced to approach survival probability in a stochastic manner. Taken together, these data further supported the initial findings (**Supplementary Table 1, Supplementary Figure 1**).

Overall immunomodulatory treatments were well tolerated. All the recorded severe adverse events were related to severe COVID-19 disease (**Supplementary Table 2**). There were no serious infections associated with the immunomodulatory therapies.

### 3.2 Disease-dependent biomarkers

There is sufficient evidence showing that inflammatory and hematological markers such as the absolute neutrophil and lymphocyte count (ANC, ALC), LDH, CRP and D-dimers, are associated with the progression and severity of COVID-19 [20–22]. As a result, we recorded these markers on the first day of progression to SRF, as well as on day 7 after the initiation of each treatment. Comparison between groups, showed statistically significant results in favor of the COMBI group, in the increase of ALC count (p=0.021) and the reduction of CRP levels (p=0.002) on day 7 (**Figure 3A**,**B**). There was no statistically significant difference between groups, regarding ANC, LDH and D-dimers values.

**Figure 3.**
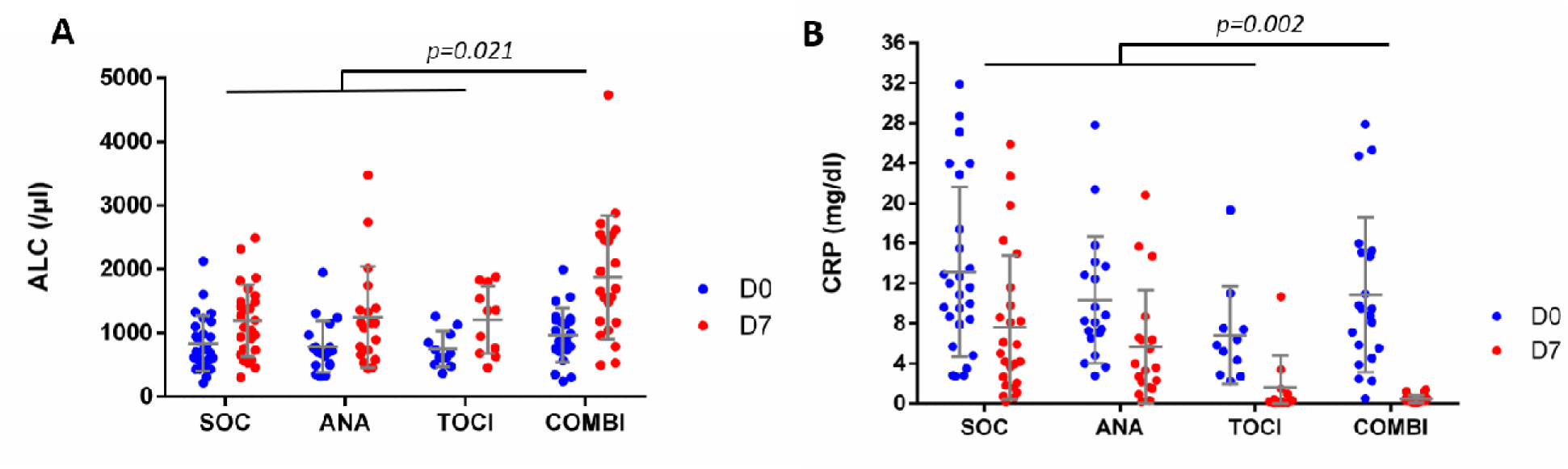
COMBI treatment **(A)** increased Absolute Lymphocyte Count (ALC) and **(B)** diminished CRP when compared with other treatments; comparisons were made between the first day of progression to SRF (D0) and the day 7 (D7) after the initiation of each treatment.

### 3.3 Primary lung fibroblasts, in vitro treated in COVID-19 environment, are involved in TF expression

Since immunothrombosis is crucially involved in pathology of COVID-19 ARDS [23], mesenchymal cell/fibroblast accumulation in lung is linked with the progression of COVID-19 severe respiratory failure [8] and fibroblasts under certain inflammatory conditions express TF [24,25], we examined whether COVID-19 environment could activate TF/thrombin pathway in LFs. We observed that plasma samples from treatment-naïve COVID-19 patients markedly induced TF expression in LFs, compared to untreated cells, as indicated by TF real-time quantitative PCR (qPCR), in-cell ELISA and immunofluorescence microscopy (**Figure 4A**,**B and D**). TF released by plasma-stimulated LFs was bioactive, as assessed by TF activity quantitative assay (**Figure 4C**). Together, our findings suggest that COVID-19 inflammatory microenvironment is a potent activator of the thrombotic potential of LFs.

**Figure 4.**
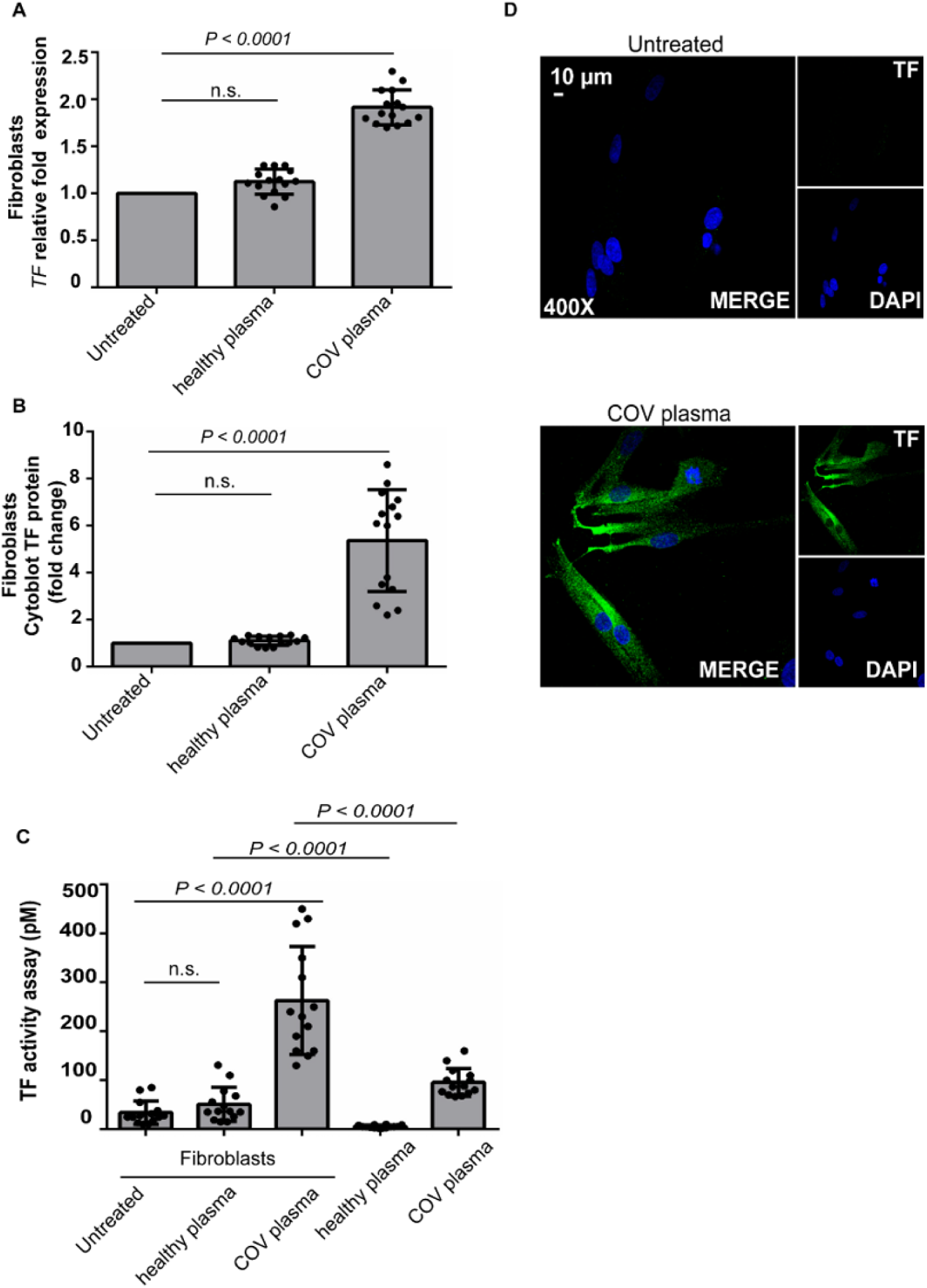
COVID-19 plasma triggers lung fibroblasts to produce tissue factor (TF) *in vitro*. Lung fibroblasts (LFs) were stimulated with 2% plasma, derived either from healthy subjects (healthy plasma) or COVID-19 patients (COV plasma). Tissue factor (TF) expression in LFs as assessed by (**A**) qPCR and (**B**) In-Cell ELISA (Cytoblot). All conditions were compared to untreated cells. (**C**) TF activity was evaluated in culture supernatants of LFs as well, in COVID-19 plasma samples as control, diluted directly with the culture medium in a final dilution that used in experimental conditions. In (**A**)-(**C**), n=15, bars represent mean±SD, (n.s.: not significant); in case of (**C**), alpha was set to 0.0125 after Bonferroni correction. (**D**) Confocal fluorescence microscopy showing TF in LFs (green: TF, blue: DAPI). A representative example of three independent experiments is shown.

### 3.4 Agents of COMBI protocol have inhibitory effect on TF/Thrombin axis in fibroblasts treated with COVID-19 plasma

According to previous findings, and considering that the proteases of TF/thrombin axis have a multivalent function leading to thrombosis, amplification of inflammation, cell proliferation, and fibrosis [25–27], we prompted to study the *in vitro* effect of the applied therapeutic regimens.

SARS-CoV-2 infection seems to activate the NF-kB signaling pathway, which may subsequently induce the secretion of multiple inflammatory cytokines, including interleukin (IL)-1, interleukin (IL)-6 and tumor necrosis factor-α (TNF-α) [28]. Since both IL-1 and IL-6 are elevated in patients with COVID-19 [14,15,29], LFs were pretreated with either a recombinant human IL-1 receptor antagonist (anakinra) or an anti-IL-6 receptor monoclonal antibody (tocilizumab), in an attempt to disrupt the autoinflammatory loops driven by these cytokines. However, TF expression (**Figure 5A-B, Supplementary Figure 2A-D**) and activity (**Figure 5C**) were not significantly attenuated in COVID plasma-stimulated LFs upon these *in vitro* inhibitions.

**Figure 5.**
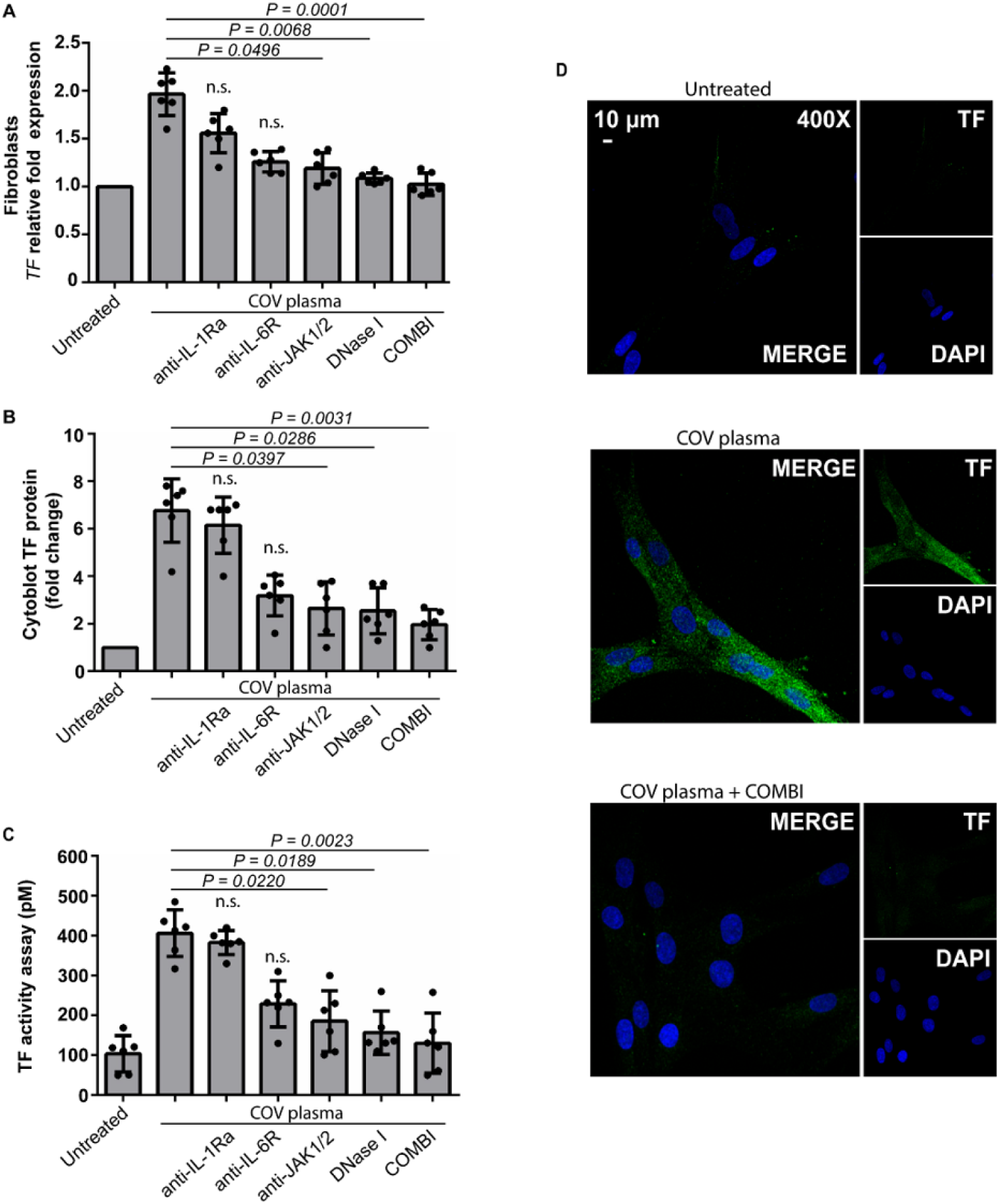
Agents of combined treatment result in reduction of tissue factor (TF) expression and activity in cultures of lung fibroblasts. Relative fold expression of (**A**) tissue factor (TF) mRNA and (**B**) In-Cell TF ELISA (Cytoblot) in lung fibroblasts (LFs) treated with 2% COVID-19-derived plasma (COV plasma) and inhibited with a recombinant IL-1 receptor antagonist (anakinra), an anti-IL-6 receptor monoclonal antibody (tocilizumab), a selective JAK1/JAK2 inhibitor (baricitinib), DNase I or combination of therapeutic agents (tocilizumab, baricitinib and DNase I). (**C**) TF activity in cell supernatants in conditions as previously described. In (**A**)-(**C**), the effect of therapeutic agents was compared to COV plasma condition, n=6, bars represent mean±SD, statistical significance was set at p<0.05 (n.s.: not significant). (**D**) Confocal fluorescence microscopy showing TF staining in stimulation and inhibition studies of LFs (green: TF, blue: DAPI). A representative example of three independent experiments is shown.

On the other hand, pre-incubation of cells with a selective JAK1/JAK2 inhibitor with an immunosuppressive role (baricitinib) [16], or pre-treatment of plasma samples with DNase I, an enzyme able to dismantle NETs [17,18], markedly reduced TF expression (**Figure 5A-B, Supplementary Figure 2E-F**) and activity (**Figure 5C**) in plasma-stimulated LFs.

In view of these data and considering that COVID-19 is mediated by plenty of plausible mechanisms, cells were simultaneously blocked against IL-6, JAK1/JAK-2 and NET signaling, leading to TF suppression in plasma-stimulated LFs (**Figure 5A-D**).

Collectively, both the separate use of DNase or JAK1/JAK2 inhibitor and the concomitant treatment of LFs with an IL-6 inhibitor, a JAK1/JAK2 inhibitor and DNase, namely the agents of COMBI protocol, could disrupt multiple sources triggering TF release from LFs, broadly preventing thromboinflammatory responses in COVID-19.

## 4. Discussion

COVID-19 patients with SRF, are in immediate need of therapeutic solutions to avoid mechanical ventilation or death. This study focused on that difficult group of patients, establishing hard inclusion criteria and endpoints. Considering the involvement of multiple mechanisms in COVID-19 pathology, it is a rational approach to apply combined therapeutic strategies aiming at reducing mortality. In this context, keeping dexamethasone and heparin as SOC treatment, we observed benefit regarding the final outcome, namely in-hospital mortality, in SRF patients by adding inhaled DNase combined with agents against IL-6 and JAK1/2 signaling. Moreover, treatment of primary lung fibroblasts, main players during the progression of respiratory failure [8,30], with the administrated immunomodulatory agents upon stimulation with COVID-19 environment, disrupted TF/thrombin expression, providing potential mechanistic insights regarding their therapeutic effect.

After the infection of respiratory epithelial cells by SARS-CoV-2, the sequence of events regarding the cross-talk of the involved inflammatory mediators, remains an obscure scientific field. A large amount of evidence has suggested the triggering of various amplification loops in a vicious cycle pattern. It is highlighted that viral proteins trigger the activation of NFκB pathway [31] and complement [11,32]. The activation of NFκB, induces the secretion of multiple chemokines and cytokines such as IL-1, IL-6, IL-8 and TNFα. Cytokine-mediated amplification loops enhance this pathway, leading to a true “cytokine storm” and influx of macrophages and neutrophils damaging the lung tissue [28]. Additionally, mediators of the inflammatory environment of COVID 19, such as, complement anaphylatoxins, platelet-derived factors, thrombin, IL-8 and G-CSF, trigger the NET-driven thromboinflammation [23,32–34]. Into the NET structures, proteases and cytokines are preserved in their active form, further amplifying thrombotic tendency, inflammation, and fibrosis [11,35,36]. Moreover, IL-6 triggers JAK/STAT signaling, driving the transcription of an extensive range of acute phase proteins such as CRP. Furthermore, IL-6 may activate and promote differentiation or proliferation of several non-immune cells, including fibroblasts [37,38]. The application of combined therapeutic regimens against COVID-19-related SRF is an emerging challenge against this complex and heterogeneous mechanistic basis of COVID-19 pathophysiology.

Heparin, as a part of SOC regimen, by potentiating antithrombin III (ATIII) activity, inhibits the hypercoagulable condition and the generation of fibrin microthrombi in the lung [39,40]. Of note, in COVID-19 low circulating levels of ATIII have been described, making the potency of heparin in these cases defective [41–44]. However, heparin may also exert its beneficial effects against immunothrombosis by dismantling thrombogenic NETs [45]. In the same context, the other partner of SOC, dexamethasone, the first agent proved to reduce mortality in a RCT [5], has anti-inflammatory/anti-cytokine effects contributing to the control of NFκB-induced hyperinflammation [46]. Recently it has been suggested that dexamethasone, by modulating COVID-19 immature neutrophils, favors the immune response [47]. In this study, we initially added on SOC, immunomodulating monotherapies using anakinra or tocilizumab consecutively.

Anakinra and tocilizumab have been tested in several RCTs that enrolled patients with COVID-19-related SRF. CORIMUNO-ANA-1, compared the use of anakinra to usual care in 116 hospitalized patients who were hypoxemic but did not require high-flow oxygen or ventilation, was stopped early for futility [48]. REMAP-CAP found that anakinra was not effective in reducing the combined endpoint of in-hospital mortality and days of organ support in ICU patients with COVID-19 [49]. Recent results of the RECOVERY and REMAP-CAP trials provided evidence that tocilizumab, when administered with corticosteroids, offers a modest mortality benefit in certain patients (including ICU patients) with COVID-19 who are severely ill, who are rapidly deteriorating and have increasing oxygen needs, and who have a significant inflammatory response [49,50].

In this study that recruited the most severely affected patients (PaO2/FiO2 <100 mmHg), the addition of anakinra or tocilizumab on top of SOC did not add significant benefit regarding mortality, need for IMV and time to discharge. As the mortality rates observed remained high, we considered that the mechanisms involved in the inflammatory environment of COVID-19 are probably interdependent and create a vicious cycle of positive feedback.

In the next, (fourth), consecutive therapeutic protocol, we added on SOC treatment a combined immunomodulation scheme, which comprised inhaled DNase to dismantle NETs and locally suppress the activity of their proteins, tocilizumab to inhibit the signalling of pre-existing IL-6 load, interrupting an initial link between JAK/STAT and NFκB, and baricitinib to further prevent the JAK/STAT dependent transcription [15,16,28,35,45,51,52]. Using this protocol (COMBI), the in-hospital mortality rate of this group of COVID-19 patients with SRF was reduced to 9.1%. In line with our clinical observations, COV-BARRIER RCT recently reported an additional survival benefit of baricitinib when added to corticosteroids in hospitalized, non-IMV, patients with COVID-19 pneumonia. The effect was most pronounced in the subgroup of patients receiving high-flow oxygen or non-invasive ventilation at baseline [53]. Additionally, it has been suggested that baricitinib has antiviral activity through interference with SARS-CoV-2 endocytosis [54].

Currently, clinical data regarding the inhaled DNase in COVID-19 are very limited, coming only from two small, non-randomized, case-control trials enrolled ICU patients with ARDS [17,18]. Treatment with nebulized DNase in most of the patients was associated with improved oxygenation and outcome, especially when used earlier in the disease course [17,18]. Decreased NETs remnants in bronchoalveolar lavage fluid was also described [18]. Patients received also corticosteroids, and in some cases convalescent plasma and anticoagulation [17,18].

Administration of COMBI led to a rapid reduction of CRP and restoration of lymphopenia, both systemic biomarkers of disease severity and progression in COVID-19 [20–22]. This is in agreement with the favorable disease outcome of these patients. Furthermore, recovery of lymphocyte count probably implies a reversal of the impaired adaptive cellular immune response described in severe COVID-19 patients [55].

Reduced mortality of SRF patients in COMBI group, prompted us to investigate the in vitro inhibitory effect of applied immunomodulatory agents, namely DNase, tocilizumab, anakinra, and baricitinib in COVID-19 culture conditions. Lung fibroblasts have been found to significantly accumulate and proliferate, repopulating the damaged alveolar wall as the disease progresses, however, their functional role in COVID-19 is still elusive [9,30,56]. Recently, Rendeiro et al described the spatial landscape of COVID-19 lung pathology in COVID-19-decedents by correlating clinical and pathological variables with high-parameter imaging mass cytometry data. Fibroblasts were closely associated with alveolar type-2 cells, large thrombi, and interventions such as intubation and treatment [8]. Moreover, S and N proteins of SARS-CoV-2 are able to induce in vitro the expression of TF on human fibroblasts [57]. Taken together, it appears that fibroblasts emerge as major players of lung tissue thromboinflammation in progressive COVID-19.

We observed an inhibitory effect on TF/thrombin axis expression and activity by primary lung fibroblasts after their treatment with the above immunomodulatory agents, used separately or combined as in the COMBI therapeutic protocol. Dismantling of NETs and inactivation of the mediators they contain, as well as JAK inhibition, seem to have a central role in countering NET driven thromboinflammation. The observed clinical outcome in all groups and the improved regulation of TF activity in supernatants obtained after treatment of cells with DNase, or/and IL-6 and JAK1/2 inhibition, indicate the involvement of various inflammatory triggers and pathways in TF/thrombin axis activation. This suggests the potential value of novel strategies based on combined immunomodulation. Several studies provide consistent evidence that TF/thrombin pathway, apart from thrombosis, is involved in inflammation and fibrotic process [27,58]. Mechanistically, fibroblasts-derived extracellular vesicles/exosomes or microparticles could transfer active TF to the lung tissue environment [59–61]. According to recent studies [11,13,32], combined targeting against other involved mediators, specifically in early stages, such as complement anaphylatoxins, may further diminish the mortality rate.

In conclusion, in this study, including non-ICU patients with SRF secondary to COVID-19 pneumonia, we found that combined compassionate therapy using inhaled DNase, tocilizumab, and baricitinib on top of SOC resulted in lower mortality and intubation rate, as well as shorter hospitalization time, compared to the use of SOC alone or along with monotherapies. Inhibition of TF/thrombin axis in lung fibroblast offers a plausible mechanistic explanation for these clinical observations. These results, although in a single-center non-RCT, encourage the design of RCTs using combined immunomodulatory therapies, which can target in a multilevel manner interconnected mechanisms of COVID-19 pathogenesis, in patients with the highest disease severity, before they cross the ICU road.

## Data Availability

All data produced in the present work are contained in the manuscript

