## Supplementary Material for "Combined administration of inhaled DNase, baricitinib and tocilizumab as rescue treatment in COVID-19 patients with severe respiratory failure"

### Supplementary Methods

#### Isolation and culture of human lung fibroblasts

Primary human lung fibroblasts (LFs) were isolated from lung lobectomy samples derived from two donors at the Academic Hospital of Alexandrouplis, Greece, as previously described [1,2]. LFs were cultured in Dulbecco's modified Eagle's medium (DMEM; Thermo Fisher Scientific, Carlsbad, SA, USA) supplemented with 10% fetal bovine serum (FBS; Thermo Fisher Scientific) and antibiotic/antimycotic solution (Thermo Fisher Scientific), at 5% CO2 and at 37oC. All experiments were carried out on fibroblast from passages 2–4. For LFs characterization, cells were stained using mouse monoclonal antibodies against alpha-smooth muscle actin (α-SMA), vimentin, and desmin (Invitrogen, Carlsbad, SA, USA), and as previously described [1].

#### Plasma collection

To isolate plasma, venous blood was collected in BD Vacutainer EDTA tubes (Becton, Dickinson and Company, Franklin Lakes, New Jersey, USA). Blood was centrifuged at 500g for 15 minutes, and then plasma samples were stored at –80°C, until analyzed [3].

#### Stimulation and inhibition studies in LFs

LFs were stimulated with EDTA plasma derived from either COVID-19 treatment-naive patients or healthy donors, at a final concentration of 2% in DMEM. To inhibit interleukin-1 (IL-1) signaling, cells were pre-treated with anakinra – an interleukin 1 receptor antagonist protein (100 ng/mL; Kineret, Orphan BIOvitrum), for 60 min. To block IL-6 signaling, cells were pre-incubated with tocilizumab – a humanized monoclonal antibody against interleukin-

6 receptor (1 ug/mL; Actemra, Hoffmann-La Roche), for 60 min. To prevent janus kinase signaling (JAK1 and JAK-2), cells were pre-treated with baricitinib (2.5 nM; Olumiant, Lilly), for 60 min. To dismantle NET structures, plasma was pre-incubated with DNase I (1 U/mL; EN0525, Thermo Fisher Scientific), for 60 min. The concentrations and time points used to examine LFs were optimized before the experiments. All substances used in the study were endotoxin free, as determined by a Limulus amebocyte assay (E8029, Sigma-Aldrich, St Louis, MO, USA).

**RNA isolation, cDNA synthesis and qPCR**

RNA isolation and cDNA synthesis were conducted in LFs as previously described [1,2]. Real-time qPCR for Tissue Factor (TF) was performed in LFs after 3h of stimulation, based on optimization experiments. GAPDH was used as the house-keeping gene to normalize the expression levels of target genes. The following oligonucleotide primers, designed by Beacon Designer™ 4.0, were used: TF (forward: 5'-TTCQGTGTTCAAGCAGTGATTCC-3'/reverse:5'-TGATGACCACAAATACCACAGC- 3') and GAPDH (forward: 5′- GGG AAG CTT GTC ATC AAT GG–3′/ reverse: 5'- CAT CGC CCC ACT TGA TTT TG–3′). The PCR protocol includes the following steps: 52°C for 5 min; 95°C for 2 min; 35 cycles of: 95°C for 15 s and 51°C for 40 s; 52°C for 5 min; melting curve analysis. Data were analyzed using the 2-ΔΔCt mathematical model [4].

**In-cell ELISA**

In-cell ELISA was performed in confluent monolayers of LFs to measure intracellular TF expression, and as previously described [3]. LFs in 96-well microplates were stimulated with plasma for 4 hours, in the presence or absence of inhibitory agents. In all conditions, cells were fixed with 8% paraformaldehyde for 30 minutes. Blocking was performed using 2× blocking solution (ab111541, Abcam, Cambridge, UK) for 2 hours. Then, 1× permeabilization buffer was added in cells for 30 minutes. After thorough washing with PBS-1X, LFs were incubated with the primary anti-human antibody against TF (10 μg/mL; sc-59714, Santa Cruz Biotechnology Inc, Dallas, Texas, USA) at 4°C, overnight. Next, horseradish peroxidase–conjugated rabbit anti–mouse IgG (1:2000 dilution; HAF007, R&D Systems, Minneapolis, USA) was added in cells, at room temperature, for 1 hour. After washing with PBS-1X, 100 μL of TMB substrate was added till blue color development. Microplates were measured at 650 nm. The corrections were done by subtracting the signal of the wells incubated in the absence of primary antibody.

**TF activity assay**

After 4h of cell stimulation, TF activity was measured in cell supernatant [3] using Tissue Factor Human Chromogenic Activity Assay Kit (ab108906, Abcam), in accordance to the manufacturer's instructions. In brief, the assay measures the ability of TF/FVIIa to activate factor X to factor Xa. The change in absorbance of the chromophore is directly proportional to the TF enzymatic activity.

For the reliability of our results, TF activity was also evaluated directly in the plasma samples that were then used as stimuli in cell cultures. In this case, plasma samples were diluted in DMEM, at the final concentration of 2%, to resemble culture conditions. These values served as controls of the assay.

**Immunofluorescence**

LFs were cultured in chambered coverslips (Ibidi, Grafelfing, Germany) and stimulated for 4h. Fixation was performed using 4% paraformaldehyde for 30 min at room temperature. Non-specific binding sites were blocked with 6% goat serum (Invitrogen) in PBS 1x. Samples were stained using mouse anti-TF monoclonal antibody (1:200 dilution; sc-59714, monoclonal IgG1, Santa Cruz Biotechnology Inc). Following three washes with PBS 1x, a polyclonal rabbit anti-mouse Alexa Fluor 488 antibody (1:1000 dilution; A-11059, Invitrogen) was utilized as secondary antibody. DAPI (Ibidi) was used for DNA counterstaining [1]. Visualization was performed in a confocal microscope (Revolution spinning disk confocal system; Andor, Ireland) with 40xlens (Olympus UPlanSApo 40x /0.95 ∞/0.11-0.23).

### Supplementary Tables

**Supplementary Table 1.** Generalized Linear Model incorporating the primary endpoint (in-hospital mortality rate) as dependent variable; all four treatment groups as well as potential confounders were included as independent variables (factors and covariates, respectively). Optimal scaling was used to convert continuous variables to binary ones.

| **Parameter** | **P-value** |
|---|---|
| **Factors** | |
| SOC group | 1.000 |
| TOCI group | 0.097 |
| ANA group | 0.888 |
| COMBI group | 0.039 |
| **Covariates** | |
| Male sex | 0.767 |
| Comorbidities (≤1 vs >1) | 0.885 |
| BMI (≤30 vs >30 kg/m$^2$) | 0.483 |
| Age (≤61 vs >61 years) | 0.974 |
| Day at admission (≤9 vs >9) | 0.455 |

**Supplementary Table 2.** Recorded severe adverse events (SAE)

| SAE | SOC n=26 | ANA n=19 | TOCI n=11 | COMBI n=22 |
|---|---|---|---|---|
| Bleeding events* | 2 | 0 | 2 | 1 |
| Pneumomediastinum | 1† | 0 | 0 | 0 |
| Pulmonary embolism | 0 | 1† | 0 | 0 |
| **SAE per group (%)** | 3 (11.5) | 1 (5.2) | 2 (18.2) | 1 (4.5) |

*Bleeding events included gastrointestinal bleeding, abdominal and arm hematomas, in need of blood transfusion; †Non-survivors

### Supplementary Figures

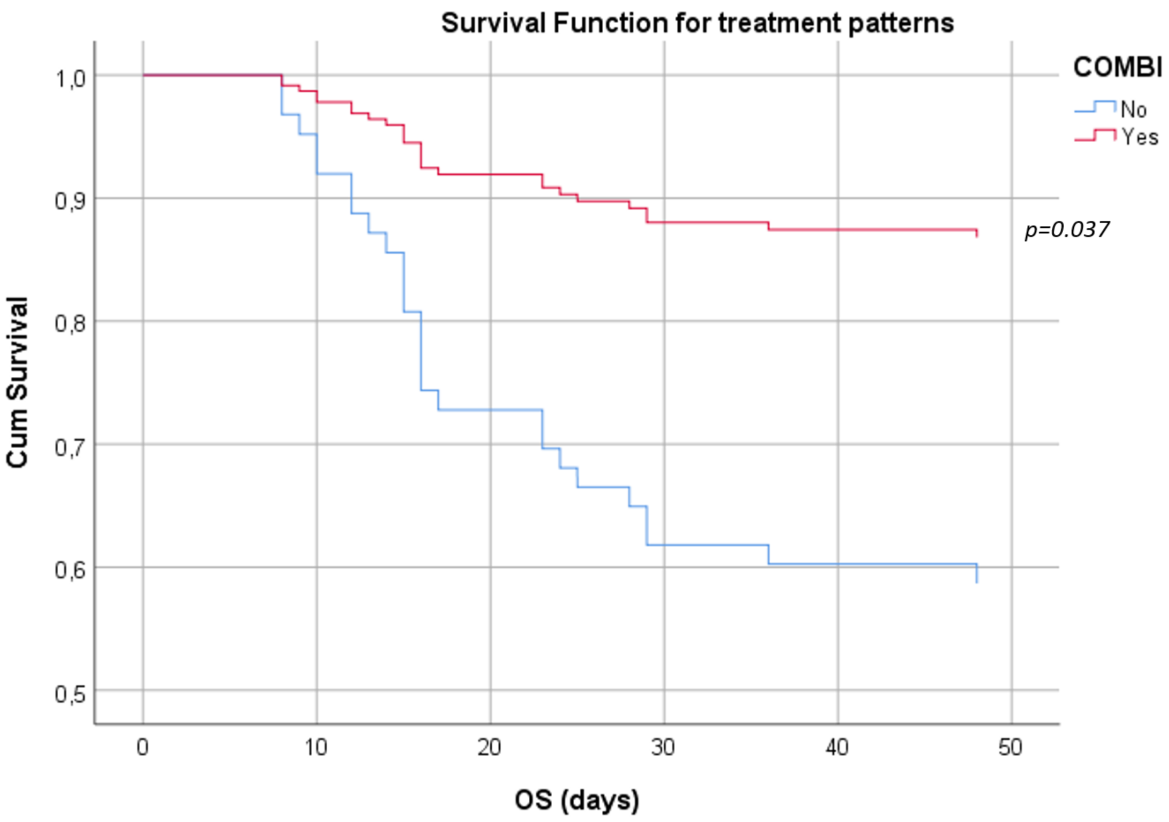


**Supplementary Figure 1.** Stochastic model based on Cox regression after adjustment for potential confounders as shown in Supplementary Table 1 [gender, comorbidities (≤1 vs >1), BMI (≤30 vs >30 kg/m$^2$), age (≤61 vs >61 years), and day at admission (≤9 vs >9)]; the administration of COMBI treatment as rescue treatment was associated with increased probability of surviving (p=0.037). Cum; cumulative, OS; overall survival.

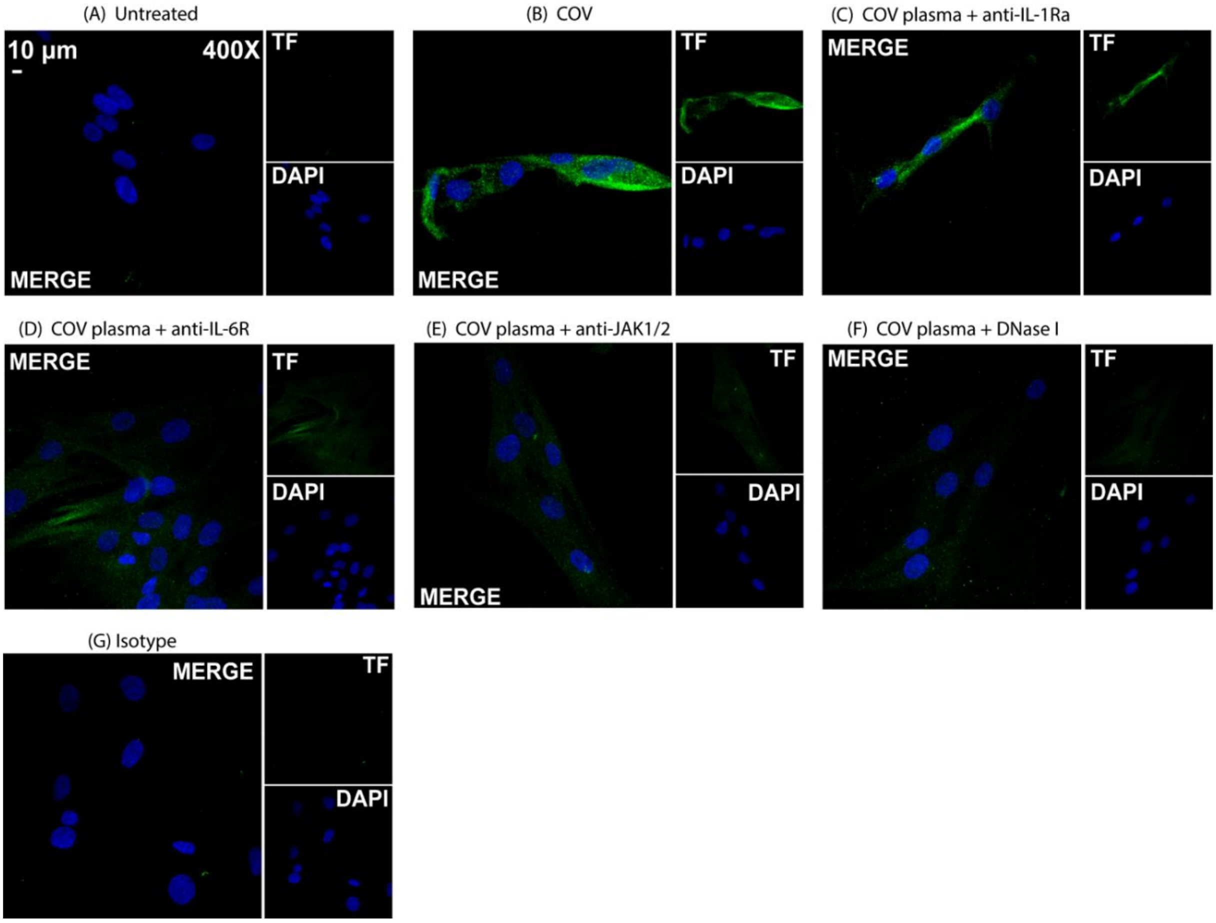


**Supplementary Figure 2. Tissue factor (TF) protein expression in human lung fibroblasts treated with plasma from COVID-19 patients.** Confocal fluorescence microscopy showing tissue factor (TF) staining in **(A)** control lung fibroblasts (LFs) **(B)** treated with 2% COVID-19-derived plasma (COV plasma) and inhibited with a **(C)** recombinant IL-1 receptor antagonist (anakinra), **(D)** an anti-IL-6 receptor monoclonal antibody (tocilizumab), **(E)** a selective JAK1/JAK2 inhibitor (baricitinib) or **(F)** DNase I. **(G)** staining with isotype control. A representative example of three independent experiments is shown (green: TF, blue: DAPI).
